# Echocardiographic Evaluation of Spironolactone on Myocardial Remodeling in Atrial Fibrillation with Preserved Ejection Fraction: the INSPIRE-AF randomized controlled trial

**DOI:** 10.1101/2023.06.08.23291178

**Authors:** Dragana Rujic, Morten Schou, Per Lav Madsen, Kenneth Egstrup

## Abstract

**Background:** By means of echocardiographic assessment and deformation analysis we sought to evaluate the effect of spironolactone versus placebo in addition to standard treatment in patients with paroxysmal or persistent atrial fibrillation (AF) and preserved ejection fraction regarding the performance of the left atrium (LA) and the left ventricle (LV), and quality of life (QOL).

**Methods:** Present double-blind, placebo-controlled study enrolled 125 patients with a history of paroxysmal (n=58) and persistent (n=67) non-valvular AF and LVEF ≥45% that were randomized to spironolactone 25 mg (n=63) or placebo (n=62) once daily in addition to optimal standard treatment. Comprehensive echocardiography and QOL were obtained at inclusion and after 12 months. The primary outcome was 12-month change in LA reservoir function as assessed by peak atrial longitudinal strain (PALS) and peak strain rate (SR-s). Secondary outcomes included LA phasic volumes, global longitudinal strain of left ventricle (GLS), E/e’ ratio, QOL, and recurrent documented episodes of AF.

**Results:** Spironolactone improved the LA reservoir function documented by PALS and SR-s (P =0.03 and P =0.02 for adjusted treatment effect, respectively) but only when adjusting for the parallel changes in blood pressure. Blood pressure significantly reduced in the spironolactone-treated subjects and affected primary outcomes, but not diastolic indices of LV. Transmitral E velocity and E/e’ ratio reduced significantly by spironolactone (P=0.009 for adjusted treatment effect). No differences in secondary outcome parameters were found across treatment groups including volumes, LA geometry, GLS, total number AF recurrences, time-to-first AF recurrence or QOL.

**Conclusion:** Spironolactone improved left atrial reservoir function by lowering blood pressure and decrease E/e’ ratio but did not affect left atrial volumes or geometry, quality of life or recurrent episodes of atrial fibrillation.

**Trial registration:** ClinicalTrials.org identifier NCT02764619 and EudraCT identifier 2013-000797-30.

## INTRODUCTION

Mineralocorticoid receptor antagonist (MRA) may have modulating effects that can attenuate and possibly even reverse adverse structural, hemodynamic, and electrophysiological maladaptive manifestations in AF (1,2). While benefits of MRA treatment are well-established in heart failure with reduced ejection fraction (HFrEF), the data in subjects without overt LV hypertrophy or systolic dysfunction, heart failure with preserved ejection fraction (HFpEF), and AF have revealed overall neutral or at best modest results.

Remodeling in AF is characterized by increased content of diffuse interstitial fibrosis in the wall of the left atrium (LA) and the myocardium of left ventricle (LV). It is a consequence to cardiac stressors that over time induce ultrastructural changes, often followed by functional and geometrical changes along with conduction heterogenicity (3,4). Extensive research emphasizes the physiological coupling of LA and LV, and in AF setting have shown a correlation between increased LA pressure and LV stiffness and atrial low-voltage areas indicative of LA arrhythmogenic substrate and poor ablation outcomes (5). A noninvasive method to display such mechanistic and functional properties is deformation analysis by speckle tracking. Speckle tracking was documented to quantify subtle changes in the myocardium, thereby providing incremental mechanistic insight before evident structural changes (6) and has contributed with predictive information in heart failure, hypertension and AF beyond that of conventional measures of LA volumes and LVEF and is increasingly accepted as an important adjunct to standard volumetric measures (7).

The INSPIRE-AF (INhibition of aldoSterone REverse remodeling in Atrial Fibrillation) trial was designed to evaluate the effect of spironolactone versus placebo in addition to standard treatment in patients with paroxysmal or persistent AF and preserved LVEF by means of echocardiographic derived indices of LA function (deformation analysis by speckle tracking) and LV diastolic function (E velocity, tissue e’, and E/e’ ratio).

## METHODS

The randomized, double-blind, placebo-controlled trial was conducted in an outpatient setting at Odense University Hospital, Svendborg, Denmark to evaluate the intervention with spironolactone 25 mg versus placebo once daily for 12 months as add-on to standard, optimal treatment of AF. Permitted background medical therapy at baseline included anticoagulants, rate control agents and treatment for coexisting conditions such as hypertension and diabetes following contemporary clinical guidelines.

Outpatients with paroxysmal or persistent AF between 2013 and 2017 were checked for eligibility by electronic medical records, 315 patients meet all inclusion criteria, 167 of whom declined participation, and 148 of whom underwent screening by transthoracic echocardiography and venous blood chemistry for serum creatinine and potassium levels. In total 125 eligible patients underwent randomization to spironolactone or placebo in a ratio 1:1 with a random block size of 2-4 and stratified by baseline type AF (paroxysmal or persistent AF) (**Figure 1**). The randomization list and study medication were organized by Odense University Hospital Pharmacy, Odense, Denmark, and the list was kept sealed behind two closed doors until time of unblinding. Enrollees, investigators, care providers and data analysts involved in screening, data management, and imaging analyses were blinded to randomization allocation.

**Figure 1.**
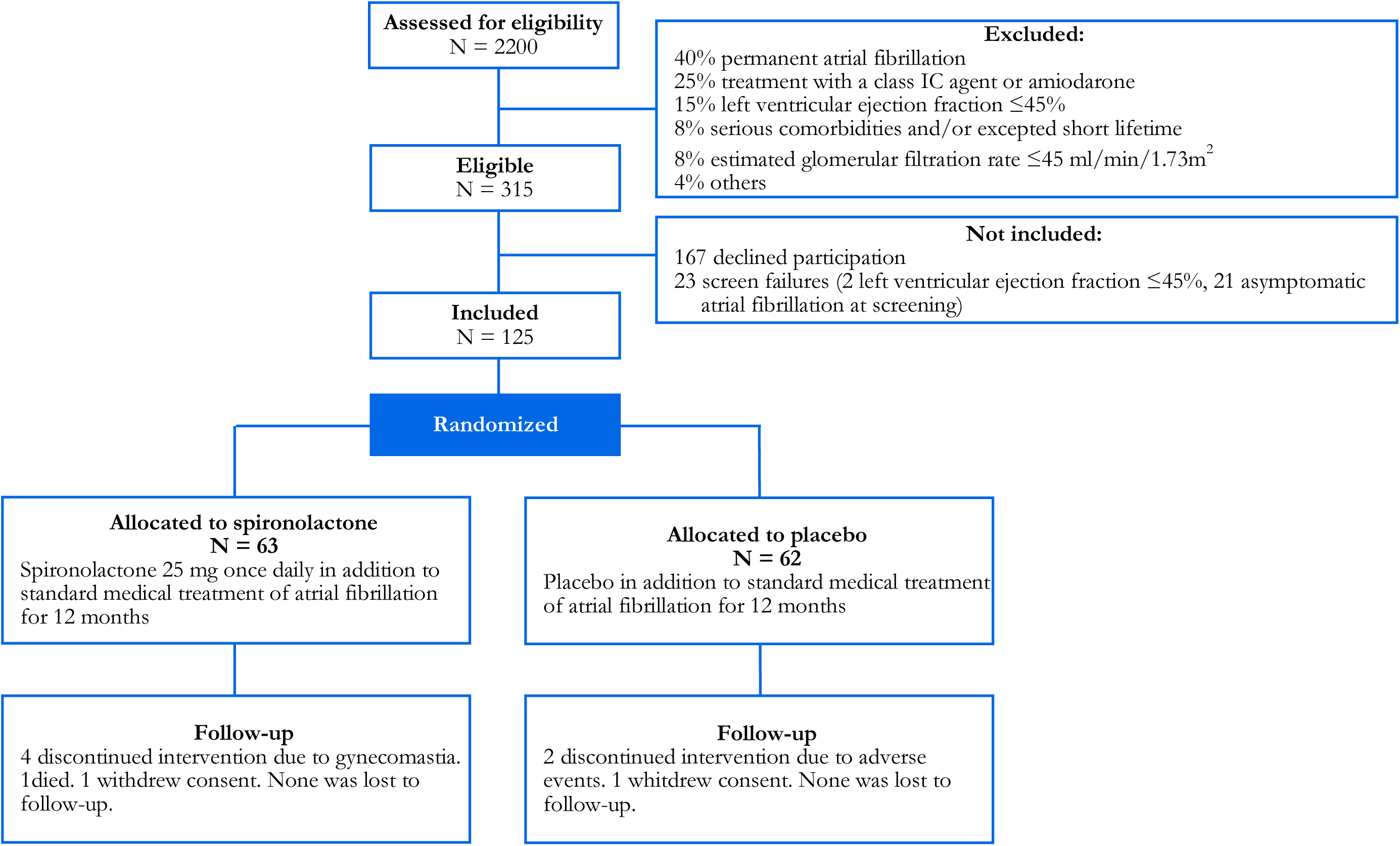
Study flow diagram

The protocol was approved by the local Ethics Committee (trial identifier S-20120220), and the Danish Medicines Agency, and was conducted in accordance with the Declarations of Helsinki and International Conference on Harmonization Guidelines for Good Clinical Practice.

### Study population

Adults with written informed consent, symptomatic paroxysmal or persistent recurrent episodes of AF classified by European Heart Rhythm Association class ≥II, in sinus rhythm at the time of randomization, at least one documented AF episode (by 12-lead ECG or AF-episode lasting >30 seconds) within previous 12 months were eligible. The main exclusion criteria were: permanent AF, previous radiofrequency or surgical treatment of AF, heart failure (New York Heart Association functional class ≥III and/or LVEF less than 45%), acute myocardial infarction within 6 months, significant valve disease, ongoing treatment with antiarrhythmic drugs (class IC or amiodarone), chronic renal dysfunction (estimated glomerular filtration rate (eGFR) ≤45 ml/min/1.73m^2^) or serum potassium ≥5.0 mmol/L. A comprehensive list of in- and exclusion criteria is provided in Supplemental **Table S1**.

### Study protocol

At baseline, prior medical history and concomitant therapy was registered, measurements of vital signs (including office blood pressure, heart rate), body weight, height, ECG, and blood chemistry (complete blood count, renal function, and electrolytes) were obtained. Follow-up visits were scheduled 1, 3, 6 and 12 months after randomization for the evaluation of clinical status, compliance with the study medication by collection of excess study medicine, vital signs, ECG, and blood chemistry. Additional blood chemistry and ECG were scheduled (and whenever clinically indicated). Timeline of study procedures and follow-up assessments are listed in Supplemental **Table S2**.

Recurrence of AF were registered during follow-up and participants were encouraged to seek a health care provider or the outpatient clinic for electrocardiographic evaluation whenever experiencing symptoms suggestive of AF. Rhythm strategy was encouraged, hence direct current (DC)-cardioversions and short-term treatment with antiarrhythmic drugs were permitted to restore sinus rhythm. In hypertensive patients, office blood pressure was controlled to achieve a level <135/85 mmHg following established clinical guidelines.

The indications for down-titration or withdrawal of study medication are described in detail in Supplemental **Table S3**. All enrollees were followed with scheduled visits and prespecified outcomes for 12 months, regardless their compliance to study medication unless withdrawal of the consent.

### Echocardiographic assessment

Standard echocardiography was carried out by Vivid E9 Digital Ultrasound equipment (GE Medical Systems, Horten, Norway) with phased array 2.5 MHz transducer, conducted by an experienced reader. Three consecutive cardiac cycles in cine loop were recorded for offline analyses adhering to the American Society of Echocardiography and the European Association of Cardiovascular Imaging (8) recommendations for assessment of following measures: LVEF using the modified biplane Simpson method, LV end-diastolic volume, LA maximal (before the opening of the mitral valve at the end of LV systole), minimal (after the closure of the mitral valve at the end of LV diastole) and pre-contract (the onset of the p-wave at the end of LV diastole) volumes using biplane area-length method, averaging apical 2- and 4-chamber values. LV diastolic function was obtained by pulsed wave Doppler from the apical 4-chamber view, including early (E) and late (A) diastolic filling velocities, and E/A ratio. The LV early diastolic tissue velocity (lateral e’) was acquired by tissue Doppler.

A semi-automated 2-dimensional speckle tracking (EchoPAC, GE Medical Systems, Horten, Norway) was applied in patients with sinus rhythm by manual tracing endocardial border of LA and endo- and epicardial border of LV in apical 2- and 4-chamber views. The region of interest was automatically computed into six segments, generating a deformation curve using the onset of QRS complex as the referent point. Manual adjustments were applied when necessary and if failed on three attempts, segments with poor tracing were excluded from further analysis. A loop was excluded if more than 2 segment dropouts, lack of full cardiac cycle or a missing view. The deformation curves were examined for peak values: peak atrial longitudinal strain (PALS) and systolic atrial strain rate (SR-s) representing LA reservoir function, early diastolic atrial SR (SR-e) represent the conduit function, peak atrial contraction strain (PACS) and late diastolic atrial SR (SR-l) representing LA active function (**Figure 2**). LV deformation was assessed, using global longitudinal strain (GLS) as an average of all LV segments interrogated.

**Figure 2.**
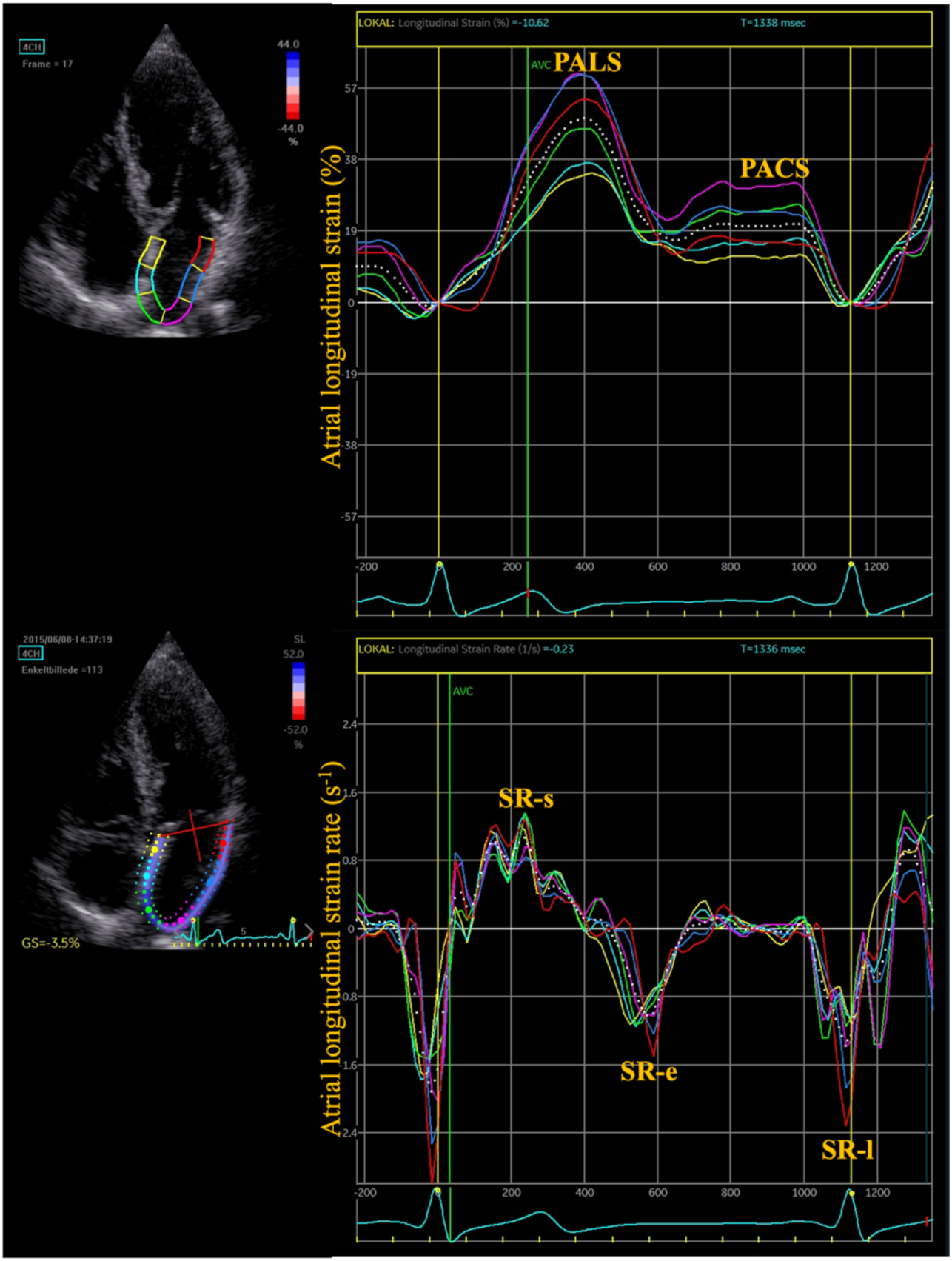
Left atrial longitudinal strain and strain rates curves during sinus rhythm. PALS indicates peak atrial longitudinal strain (PALS); PACS, peak atrial contraction strain; SR-s, systolic atrial strain rate, SR-e, early diastolic atrial strain rate; and SR-l, late diastolic atrial strain rate.

Three enrollees did not undergo follow-up scan, and a total of 124 and 123 echocardiographic studies were available for quantitative analysis in the spironolactone and placebo group, respectively. LA deformation analyses were possible equally in 112 scans (87.5% vs. 89.3%) in the spironolactone and placebo group, respectively. A total of 20 randomly selected studies underwent repeat review for assessment of inter- and intra-rater variability of the primary endpoint (Supplemental **Table S4**).

### Quality of life

Quality of life (QOL) was evaluated at baseline and at 12-month follow-up by 12-Item Short-Form Health Survey (SF-12) questionnaire and a structured interview by the Modified Toronto Atrial Fibrillation Severity Scale (mAFSS). SF-12 answers were entered in dedicated software (QualityMetric Health Outcomes Scoring System 5.0, US), generating a mean physical and mental summary score with a range 0-100 points, with lower scores reflecting poorer QOL (9). At scheduled visits subjects were interviewed using the mAFSS algorithm to assess severity of AF symptoms regarding information of frequency, duration, and severity of AF symptoms; each quantity ranging 3-30 contributing equally to the total AF burden score, and higher scores reflecting worse AF symptoms (10,11).

### Statistical analysis

Continuous variables were described as mean ±standard deviation and between groups comparisons were made by Mann-Whitney U test or Student’s t test, as appropriate. Categorical variables were described as frequency and percentage, and comparisons were made by Pearson’s chi-square test. Changes in longitudinal data, including vital signs, laboratory measurements, and treatment effects of echocardiographic primary and secondary outcomes, were obtained by mixed linear models and missing data were handled by maximized likelihood. As indices of LA function such as volumes, strain and strain rate measurements are load-dependent, the changes in prespecified parameters, including body mass index, systolic and diastolic blood pressure, were applied as covariates in the multiple linear regression models as they were assumed to change during the intervention. Time-to-first events analyses were carried out using Cox proportional hazards model with prespecified baseline covariates sex, age, type of AF, and blood pressure. Differences between time-to-event curves were compared with log-rank test. For the analysis of total number (first and repeated) events, the differences between treatment groups were assessed by means of binomial negative model. Only ECG documented AF episodes were included in the statistical analyses.

All statistical analyses were carried out on intention-to-treat basis, including all randomized subjects. No imputations were made. Statistical significance was defined as p<0.05. Stata version 16 (Texas, US) was used for all analyses.

### Outcomes

The primary endpoint was 12-month change in the reservoir function of LA. Sample size calculations are described in Supplemental material.

Secondary imaging outcomes included LA phasic volumes and E/e’. Non-imaging secondary outcomes included QOL assessed by SF-12 questionnaire and structured interview mAFSS, first AF recurrence and DC cardioversion analyzed as time-to-first event, secondly all AF recurrences and DC cardioversions (including first and repeated events). Furthermore, prespecified safety outcomes including gynecomastia, hyperkalemia and renal impairment were assessed.

## RESULTS

### Patient selection and characteristics

The study ended according to protocol. Only 125 patients were recruited to the study compared to anticipated 130 patients. However, none were lost to follow-up and only 3 patients did not complete the 12 months follow-up; one patient allocated to the spironolactone group and one patient allocated to the placebo group withdrew consent (2 and 140 days after the randomization, respectively), finally one diabetic patient allocated to the spironolactone group died of alcohol intoxication (184 days after the randomization). With respect to the sample size calculations, we therefore consider our results to be valid.

The allocated groups were well-balanced with respect to demographics, vital signs, and prevalence of comorbidities (**Table 1**); the mean age was 63.9±8.1 years; 64% were male; 46.4% had paroxysmal AF; and 53.6% had persistent AF. Comorbid hypertension was common, 51.6% of all patients; 32% of whom received angiotensin-converting-enzyme inhibitor (ACEI) or angiotensin II-receptor blocker (ARB).

**Table 1.**
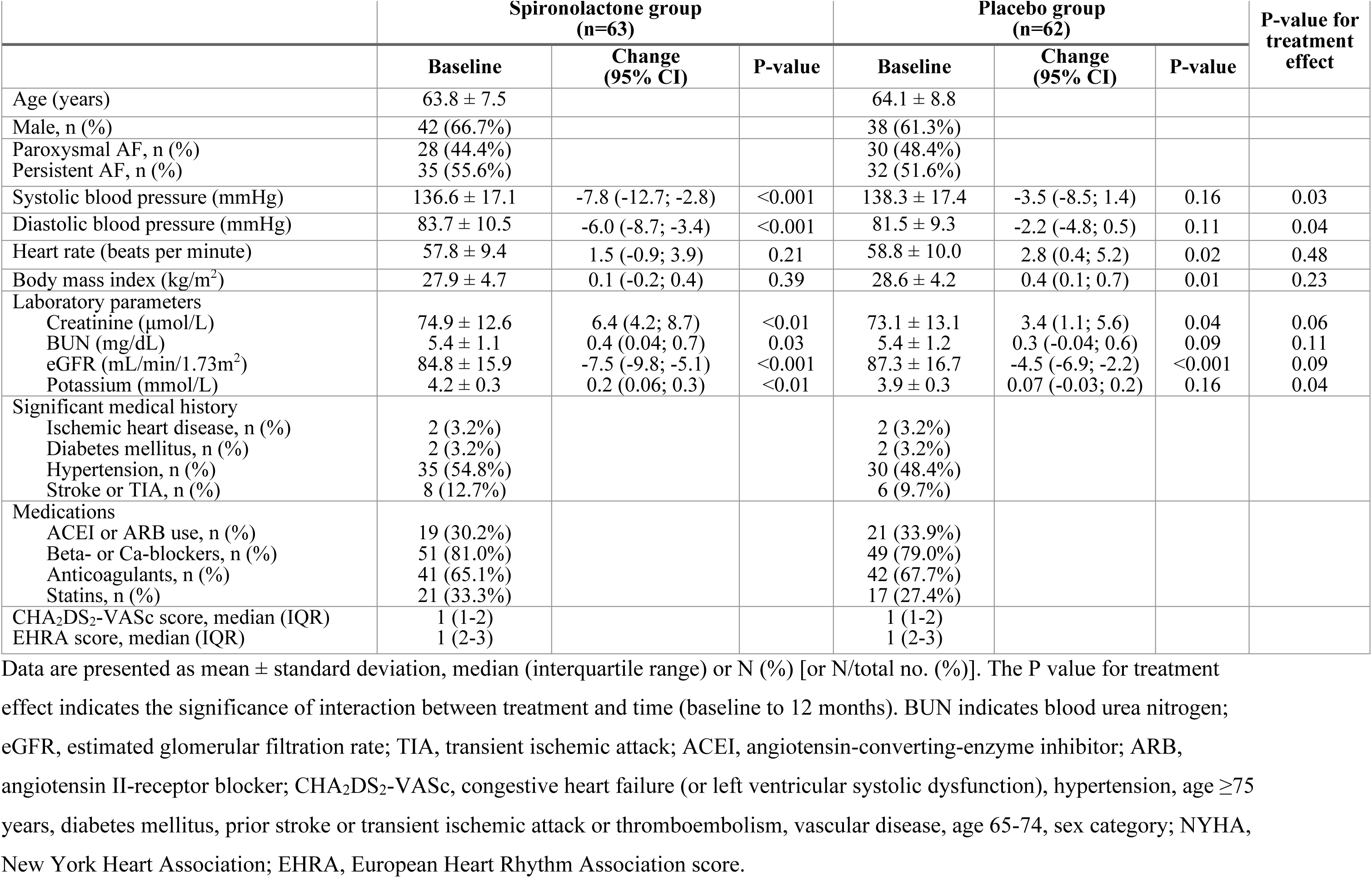
Clinical and demographic characteristics, and 12-month change in clinical characteristics

|  | Spironolactone group<br>(n=63) |  |  | Placebo group<br>(n=62) |  |  | P-value for<br>treatment<br>effect |
| --- | --- | --- | --- | --- | --- | --- | --- |
|  | Baseline | Change<br>(95% CI) | P-value | Baseline | Change<br>(95% CI) | P-value |  |
| Age (years) | 63.8 ± 7.5 |  |  | 64.1 ± 8.8 |  |  |  |
| Male, n (%) | 42 (66.7%) |  |  | 38 (61.3%) |  |  |  |
| Paroxysmal AF, n (%) | 28 (44.4%) |  |  | 30 (48.4%) |  |  |  |
| Persistent AF, n (%) | 35 (55.6%) |  |  | 32 (51.6%) |  |  |  |
| Systolic blood pressure (mmHg) | 136.6 ± 17.1 | -7.8 (-12.7; -2.8) | <0.001 | 138.3 ± 17.4 | -3.5 (-8.5; 1.4) | 0.16 | 0.03 |
| Diastolic blood pressure (mmHg) | 83.7 ± 10.5 | -6.0 (-8.7; -3.4) | <0.001 | 81.5 ± 9.3 | -2.2 (-4.8; 0.5) | 0.11 | 0.04 |
| Heart rate (beats per minute) | 57.8 ± 9.4 | 1.5 (-0.9; 3.9) | 0.21 | 58.8 ± 10.0 | 2.8 (0.4; 5.2) | 0.02 | 0.48 |
| Body mass index (kg/m<sup>2</sup>) | 27.9 ± 4.7 | 0.1 (-0.2; 0.4) | 0.39 | 28.6 ± 4.2 | 0.4 (0.1; 0.7) | 0.01 | 0.23 |
| Laboratory parameters |  |  |  |  |  |  |  |
| Creatinine (μmol/L) | 74.9 ± 12.6 | 6.4 (4.2; 8.7) | <0.01 | 73.1 ± 13.1 | 3.4 (1.1; 5.6) | 0.04 | 0.06 |
| BUN (mg/dL) | 5.4 ± 1.1 | 0.4 (0.04; 0.7) | 0.03 | 5.4 ± 1.2 | 0.3 (-0.04; 0.6) | 0.09 | 0.11 |
| eGFR (mL/min/1.73m<sup>2</sup>) | 84.8 ± 15.9 | -7.5 (-9.8; -5.1) | <0.001 | 87.3 ± 16.7 | -4.5 (-6.9; -2.2) | <0.001 | 0.09 |
| Potassium (mmol/L) | 4.2 ± 0.3 | 0.2 (0.06; 0.3) | <0.01 | 3.9 ± 0.3 | 0.07 (-0.03; 0.2) | 0.16 | 0.04 |
| Significant medical history |  |  |  |  |  |  |  |
| Ischemic heart disease, n (%) | 2 (3.2%) |  |  | 2 (3.2%) |  |  |  |
| Diabetes mellitus, n (%) | 2 (3.2%) |  |  | 2 (3.2%) |  |  |  |
| Hypertension, n (%) | 35 (54.8%) |  |  | 30 (48.4%) |  |  |  |
| Stroke or TIA, n (%) | 8 (12.7%) |  |  | 6 (9.7%) |  |  |  |
| Medications |  |  |  |  |  |  |  |
| ACEI or ARB use, n (%) | 19 (30.2%) |  |  | 21 (33.9%) |  |  |  |
| Beta- or Ca-blockers, n (%) | 51 (81.0%) |  |  | 49 (79.0%) |  |  |  |
| Anticoagulants, n (%) | 41 (65.1%) |  |  | 42 (67.7%) |  |  |  |
| Statins, n (%) | 21 (33.3%) |  |  | 17 (27.4%) |  |  |  |
| CHA<sub>2</sub>DS<sub>2</sub>-VASc score, median (IQR) | 1 (1-2) |  |  | 1 (1-2) |  |  |  |
| EHRA score, median (IQR) | 1 (2-3) |  |  | 1 (2-3) |  |  |  |

Data are presented as mean ± standard deviation, median (interquartile range) or N (%) [or N/total no. (%)]. The P value for treatment effect indicates the significance of interaction between treatment and time (baseline to 12 months). BUN indicates blood urea nitrogen; eGFR, estimated glomerular filtration rate; TIA, transient ischemic attack; ACEI, angiotensin-converting-enzyme inhibitor; ARB, angiotensin II-receptor blocker; CHA<sub>2</sub>DS<sub>2</sub>-VASc, congestive heart failure (or left ventricular systolic dysfunction), hypertension, age ≥75 years, diabetes mellitus, prior stroke or transient ischemic attack or thromboembolism, vascular disease, age 65-74, sex category; NYHA, New York Heart Association; EHRA, European Heart Rhythm Association score.

### Changes in blood pressure and adverse events

During the intervention a greater mean reduction in blood pressure were observed in the spironolactone group (mean reduction in the systolic blood pressure: 7.8 mmHg [95% confidence interval (CI): -12.7; -2.8] vs. 3.5 mmHg [95% CI: -8.5; 1.4] in the spironolactone vs. placebo group, respectively; P=0.03 for comparison between groups) (**Table 1**). Spironolactone treated patients also had greater mean increase in serum potassium (P=0.04 for comparison between groups) and a modest increase in serum creatinine, although the latter not reaching statistical significance. Only one patient allocated to the intervention group suffered severe hyperkalemia and a parallel increase in serum creatinine ≥30% from baseline.

Not unexpectedly, a higher occurrence of gynecomastia was noted in the spironolactone group (P=0.02 for comparison between groups); a total of 8 (12.7%) patients allocated to spironolactone group experienced this adverse event, of whom 4 (6.3%) patients prematurely discontinued the study medication (mean 202±18 days from the randomization). Full lists of adverse events are available in Supplemental **Table S5**, **S6** and **Figure S1**.

### Primary outcomes

When compared with placebo, the reservoir function of LA improved with spironolactone therapy as evidenced by a parallel increase in PALS and SR-s. In the placebo group, however, both indices of the reservoir function of LA worsened at 12-months follow-up as reflected by reduced PALS and SR-s (mean increase in PALS in spironolactone group: 1.8% [95% CI: -1.2; 4.8]; vs. mean reduction in placebo group: 3.0% [95% CI: -6; -0.07]; P =0.03 for adjusted treatment effect) (**Table 2**, **Figure 3**).

**Table 2.** Comparison of structure, function and geometry of LA at baseline and end-of-study, assessed by transthoracic echocardiography

|  | Spironolactone group<br>(n=63) |  |  | Placebo group<br>(n=62) |  |  | P-value for<br>treatment<br>effect |
| --- | --- | --- | --- | --- | --- | --- | --- |
|  | Baseline | Change<br>(95% CI) | P-value | Baseline | Change<br>(95% CI) | P-value |  |
| **LA Reservoir function** |  |  |  |  |  |  |  |
| LAVImax (mL/m<sup>2</sup>) | 35.6±11.4 | -6.9 (-17.7; 5.3) | 0.30* | 34.9±9.8 | -4.1 (-14.9; 8.1) | 0.50* | 0.70* |
| LAVImin (mL/m<sup>2</sup>) | 18.3±7.6 | -13.9 (-25.9; -0.08) | 0.049* | 17.1±6.8 | -3.2 (-16.7; 11.2) | 0.60* | 0.30* |
| LA reservoir volume index (mL/m<sup>2</sup>) | 17.4±5.4 | -2.8 (-14.8; 11.1) | 0.70* | 17.7±4.8 | -5.1 (-16.6; 8.0) | 0.40* | 0.80* |
| LA TEF (%) | 49.7±8.7 | 3.3 (-0.2; 6.8) | 0.06 | 51.8±9.2 | -0.4 (-3.7; 3.0) | 0.80 | 0.10 |
| PALS (%) | 29.0±9.2 | 1.8 (-1.2; 4.8) | 0.20 | 31.0±9.5 | -3.0 (-6.0; -0.007) | 0.05 | 0.03 |
| LA SR-s (1/s), median (IQR) | 1.1 (1-1.4) | 10.2 (0.5; 22.1) | 0.06* | 1.2 (1-1.6) | -7.0 (-15.9; 2.8) | 0.20* | 0.02* |
| **LA Conduit function** |  |  |  |  |  |  |  |
| LAVprec (mL/m<sup>2</sup>) | 25.5±10.3 | 2.9 (-10.0; 17.5) | 0.70* | 25.2±7.8 | -1.2 (-13.3; 12.6) | 0.90* | 0.70* |
| LA passive volume index (mL/m<sup>2</sup>) | 10.1±5.4 | -2.2 (-4.3; -0.1) | 0.04 | 9.7±5.8 | -1.1 (-3.2; 0.9) | 0.30 | 0.50 |
| LA PEF (%) | 29.2±13.6 | -6.9 (-11.6; -2.1) | <0.01 | 27.1±12.7 | -2.5 (-7.2; 2.1) | 0.30 | 0.10 |
| LA SR-e (1/s), median (IQR) | -1.1 (-1.3 to -0.9) | -2.1 (-17.3; 12.4) | 0.80* | -1.1 (-1.4 to -0.9) | -21.9 (-31.9; -10.7) | <0.0001* | 0.02* |
| **LA Active function** |  |  |  |  |  |  |  |
| LA active volume index (mL/m<sup>2</sup>) | 7.4±5 | 2.5 (0.4; 4.6) | 0.02 | 8.1±4.5 | 0.2 (-1.9; 2.3) | 0.80 | 0.10 |
| LA AEF (%) | 27.6±14.2 | 11.5 (6.7; 16.3) | <0.001 | 32.5±14.4 | 4.1 (-0.6; 8.6) | 0.09 | 0.02 |
| PACS (%) | 13.9±5.9 | 1.8 (-0.1; 3.6) | 0.06 | 15.9±5.9 | -0.6 (-2.4; 1.2) | 0.50 | 0.07 |
| LA SR-l (1/s), median (IQR) | -1.7 (-1.2 to -2.2) | 0.1 (-0.1; 0.3) | 0.20 | -1.7 (-1.3 to -1.7) | -0.1 (-0.3; 0.1) | 0.20 | 0.09 |
| **LA Geometry** |  |  |  |  |  |  |  |
| LA maximum length (mm) | 60.2±7.9 | 0.1 (-0.11; 0.4) | 0.30 | 60.1±7.1 | 0.2 (-0.1; 0.4) | 0.20 | 0.90 |
| LA sphericity (%), median (IQR) | 0.31 (0.23 to 0.45) | 0.2 (-0.5; 0.8) | 0.10* | 0.37 (0.23 to 0.55) | 0.4 (0.1; 1.2) | 0.003* | 0.30* |

Continuous variables are given as mean ± standard deviation (approximately normally distributed variables, i.e. LAmax, left atrial maximum volume; LAVI max, left atrial maximum volume indexed to body surface area; LAmin, left atrial minimum volume; LAVImin, left atrial minimum volume ; LApre, left atrial pre-contraction volume in mid-diastole; LApreI, left atrial pre-contraction volume indexed to body surface area; LA TEF, left atrial total emptying fraction; LA PEF, left atrial passive emptying fraction; LA AEF, left atrial active emptying fraction; reservoir, passive, and active volume; PALS, peak atrial longitudinal strain; PACS, peak atrial contraction longitudinal strain) or median (IQR) (non-normally distributed variables, i.e. variables SR-s, systolic atrial strain rate; SR-e, early diastolic atrial strain rate; SR-l, late diastolic atrial strain rate; LA sphericity).
Data are compared using mixed-effects linear models, where between subject effect is treatment and the within-subject factor is time. All models were adjusted for type AF at baseline, i.e. paroxysmal or persistent AF, age, body mass index, and change in blood pressure. The P value for treatment effect indicates the significance of interaction between treatment and time (baseline to 12 months).
\*Natural log transformation of the variable

**Figure 3.**
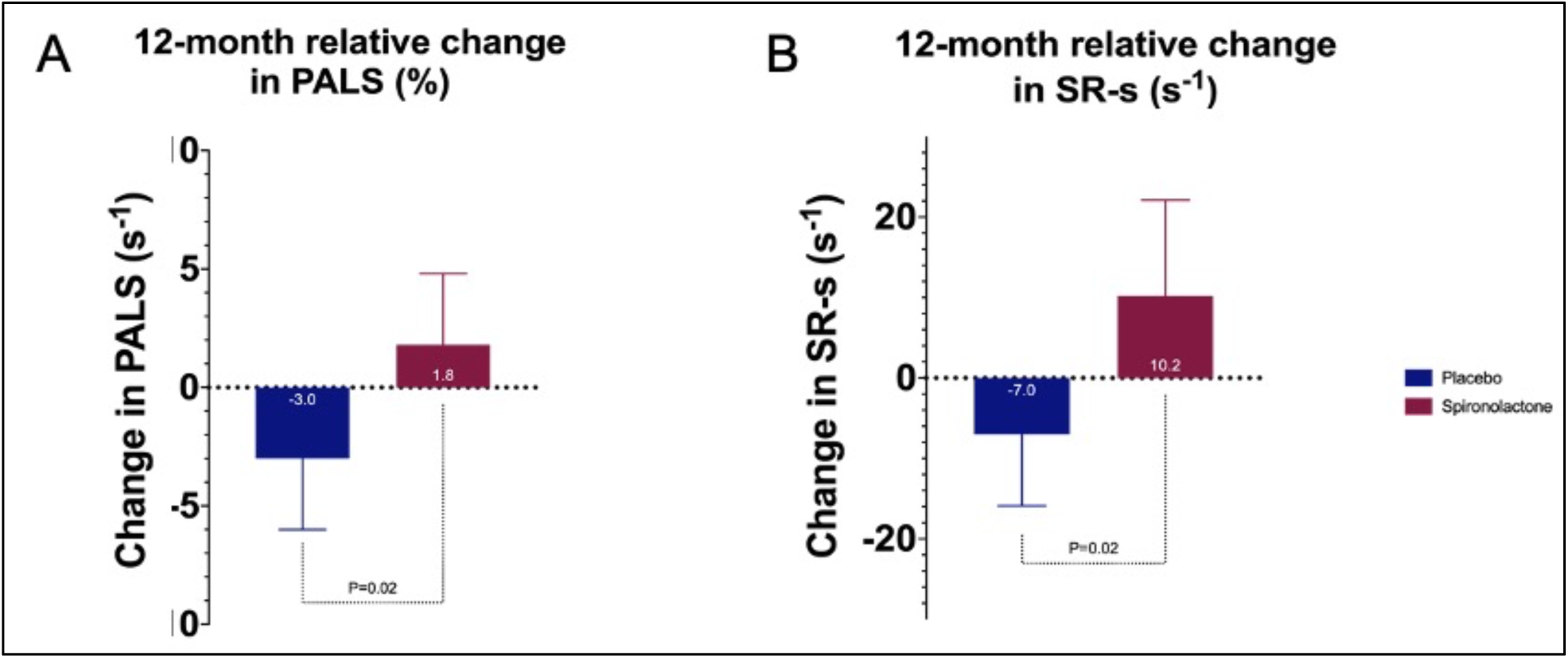
Left atrial reservoir function evaluated by deformation analysis. (A) The relative reduction in peak atrial longitudinal strain (PALS) (%) according to treatment arm in the study. The bars denote mean PALS reductions, whereas the lines represent 95% confidence intervals. (B) The relative reduction in peak systolic atrial strain rate (SR-s) (s^-1^) according to treatment arm in the study. The bars denote mean SR-s reductions, whereas the lines represent 95% confidence intervals.

The intervention group had greater absolute reduction in LA maximal and minimal volumes, but the difference to the placebo did not reach statistical significance. Indices of volume and deformation analyses during the active phase of LA increased consistently to a greater extent with the spironolactone treatment, and LA active emptying fraction was significantly improved at follow-up (P=0.02 for adjusted treatment effect) (**Table 2**). Spironolactone did not influence LA sphericity index.

The improvements of the LA reservoir function and the consistent findings across volumes in the reservoir and the active phases with spironolactone were dependent of changes in blood pressure and BMI when adjusted for in the regression models. Further exploration was performed by leaving changes in blood pressure and BMI out of the statistical analysis, and the difference between treatment groups remained significant (**Table 4**).

**Table 4.**
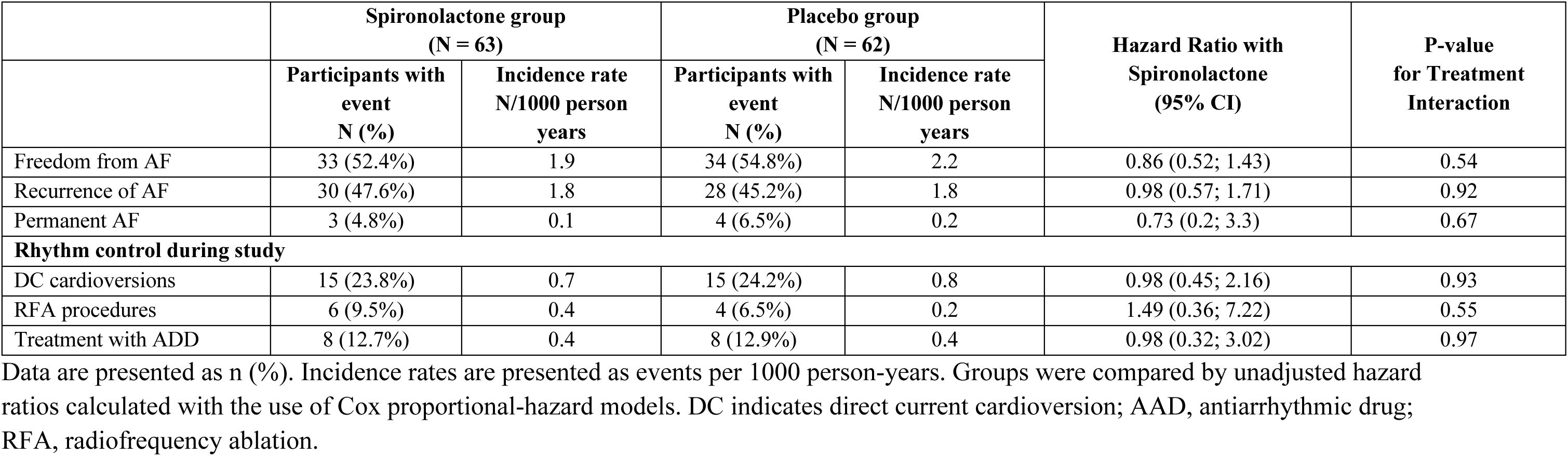

### Secondary imaging outcomes

In comparison with placebo, there were no notable changes with the active treatment in echocardiographically derived measurements of LV structure or performance (**Table 3**). There was, however, an improvement in diastolic function indexes in terms of reduction in transmitral E velocity and E/e’ ratio with spironolactone therapy, whilst both increased in placebo group (mean reduction in E/e’ ratio in spironolactone group: 0.3 [95% CI: -0.8; 0.3] vs. mean increase in E/e’ ration in placebo group: 0.8 [0.2; 1.3]; P=0.009 for adjusted treatment effect). Of special note, tissue Doppler also revealed significantly improved late diastolic annular velocity in the spironolactone group (lateral a’, P=0.03 for adjusted treatment effect).

**Table 3.** Comparison of structure, function, fibrosis burden of LV at baseline and end-of-study, assessed by transthoracic echocardiography

|  | Spironolactone group<br>(n=63) |  |  | Placebo group<br>(n=62) |  |  | P-value for<br>treatment<br>effect |
| --- | --- | --- | --- | --- | --- | --- | --- |
|  | Baseline | Change<br>(95% CI) | P-value | Baseline | Change<br>(95% CI) | P-value |  |
| **LV Structure** |  |  |  |  |  |  |  |
| LVEDVI (mL/m<sup>2</sup>) | 56.9±16.1 | -2.5 (-6.5; 1.4) | 0.20 | 56.8±13.9 | -0.3 (-4.4; 3.8) | 0.90 | 0.40 |
| LVESVI (mL/m<sup>2</sup>) | 22.9±5.3 | -3.2 (-13.6; 8.4) | 0.60* | 23.3±6.5 | 10.9 (-1.1; 24.4) | 0.08* | 0.09* |
| LVSV (mL) | 70.3±23 | -3.5 (-12.2; 5.9) | 0.50* | 69.6±19.8 | 4.4 (-5.1; 14.9) | 0.40* | 0.20* |
| LVMI (g/m<sup>2</sup>) | 76.1±17.3 | -4.3 (-9.4; 1.0) | 0.10* | 72.3±14.9 | -0.9 (-4.3; 6.3) | 0.70* | 0.20* |
| **LV Function** |  |  |  |  |  |  |  |
| LVEF (%) | 59.6±3.8 | 0.6 (-0.9; 2.2) | 0.50 | 59.6±4.1 | -0.2 (-1.8; 1.4) | 0.80 | 0.50 |
| GLS (%), median (IQR) | -19.9 (-21.6 to -18.0) | -0.7 (-1.9; 0.4) | 0.20 | -20.2 (-21.9 to -16.9) | -2.1 (-3.1; -0.9) | <0.001 | 0.10 |
| **Hemodynamics** |  |  |  |  |  |  |  |
| E velocity (m/s) | 0.68±0.2 | -0.02 (-0.06; 0.01) | 0.20 | 0.70±0.2 | 0.04 (0.003; 0.07) | 0.03 | 0.02 |
| E/A ratio, median (IQR) | 1.0 (0.8 to 1.3) | -3.2 (-12.7; 7.2) | 0.51* | 0.99 (0.9 to 1.2) | -0.5 (-10.2; 10.1) | 0.87* | 0.74* |
| Deceleration time (m/s) | 236.7±71.7 | 6.6 (-2.9; 17.1) | 0.27* | 236.1±52.9 | -0.5 (-9.3; 9.1) | 0.93* | 0.11* |
| Lateral e' (cm/s) | 0.09 ± 0.03 | -5.8 (-14.2; 3.5) | 0.18* | 0.09±0.02 | -4.4 (-12.9; 5.0) | 0.32* | 0.77* |
| Lateral a' (cm/s) | 0.08 ± 0.03 | 0.02 (0.008; 0.03) | <0.0001 | 0.08±0.02 | 0.003 (-0.005; 0.01) | 0.40 | 0.03 |
| E/e' ratio, median (IQR) | 7.2 (6.3 to 9.4) | -0.3 (-0.8; 0.3) | 0.21 | 7.6 (6.2 to 8.7) | 0.8 (0.2; 1.3) | 0.008 | 0.009 |

Data presented as mean ± standard deviation, median (IQR) or percent %. The P value for treatment effect indicates the significance of interaction between treatment and time (baseline to 12 months). LV indicates left ventricular; LVEDV, LV end-diastolic volume; LVEDVI, LVEDV index to body surface area calculated by DuBois DuBois formula; LVESVI, LV end-systolic volume index; LVMI, LV mass index; GLS, global longitudinal strain; SrL peak E, peak early diastolic longitudinal strain rate; SrL peak A, peak late diastolic longitudinal strain rate; SrL peak S, peak systolic longitudinal strain; A, late diastolic peak velocity of mitral flow; a', peak late diastolic annular velocity; E, early diastolic peak velocity of the mitral flow; e', peak early diastolic annular velocity.
\*Natural log transformation of the variable

### Changes in quality of life and recurrence of AF

From baseline to 12-months of follow-up, there were minimal mean absolute improvements in the physical and mental SF-12 scores, and similarly for the mAFSS symptom, burden and total scores in the spironolactone treated patients, but none reached statistical significance (Supplemental **Table S7**).

Consistent throughout the clinical outcomes, the treatment arms did not differ regarding sinus rhythm maintenance, number of AF recurrences and DC cardioversions during follow-up (**Table 4**). Perhaps by a chance finding and given the small event rate, patients assigned to placebo experienced fewer AF recurrences, DC cardioversions, and more patients remaining free of AF compared to the spironolactone group. However, placebo treated patients had more frequent use of transient antiarrhythmic therapy and more often suffered permanent AF. The risk of recurrent number of AF and DC cardioversions did not differ between treatment arms by means of binomial negative model revealing IRR 0.79 [95% CI: 0.57; 1.11], P=0.18.

## DISCUSSION

The principal findings provided by current study are that 12-months treatment with spironolactone in patients with preserved LVEF and adequate standard therapy of paroxysmal and persistent AF demonstrated: 1. improved LA reservoir function largely driven by a parallel reduction in blood pressure; 2. amelioration of LV diastolic properties that were independent of changes in blood pressure; 3. no changes in quality of life, recurrent episodes of AF or time-to-first recurrent AF episode.

In current AF population with minimal to no structural heart disease, while spironolactone improved the reservoir function of LA (as evidenced by increased PALS and Sr-e) and the diastolic indices of LV (as evidenced by attenuated E/e’) along with significant reduction in blood pressure, the conventional echocardiographic measurements of LA and LV volumes remained unaffected. One may speculate whether the observed positive mechanistic changes by spironolactone may be a primarily hemodynamic response rather than actual reverse-remodeling effect mitigating adverse myocardial fibrosis (12,13). Intriguingly, the mechanistic properties of LA and LV in the placebo group deteriorated over time despite no differences in baseline measures of LA and LV, proportion of patients with hypertension or differences in background concomitant medical therapy between groups. Indeed, both treatment groups achieved the ambulatory blood pressure of <135/85 mmHg after 12-months follow-up. This raises the possibility that the positive outcomes with spironolactone may not be an exclusive load-depended response. Earlier work with aldosterone antagonism revealed a correlation between reduced aldosterone levels in the circulation, downregulation of myocardial aldosterone synthesis, and ameliorated myocardial fibrosis independent of blood pressure, hence reinforcing that blood pressure is not of primary importance to the cardioprotective effects exerted by MRA (14,15). Further, MRA have a superior blockade of aldosterone synthesis, prohibiting aldosterone escape phenomenon, combined with a less antihypertensive influence compared to ACEI/ARB (16–18).

Of note, in our study, spironolactone affected PALS, but not GLS. Intervention trials with MRA in non-AF cohorts showed improved PALS and LV diastolic function by means of reduced E/e’, LV mass index, left atrial diameter along with reduced collagen turnover with greater effect in patients with higher levels of collagen metabolism (19–23). Similar our data, GLS remained unaffected by spironolactone, suggesting that PALS may be more load-depended or responsive to MRA than GLS, or that the changes in PALS and GLS do not correlate in a simple linear fashion.

In contrast to PALS, our data showed significantly reduced E/e’ ratio independent of alternations in blood pressure. Indeed, other investigations demonstrated similar improvement in LV filling pressure by MRA therapy accompanied by lowered atrial pressure, mechanistic stress and stretch of the atrial wall, leading to improved atrial compliance and the atrial reservoir function (7). Given the atrioventricular coupling, E/e’ ratio is a measure of passive/active emptying/filling between LV and LA during a cardiac cycle, accordingly the E/e’ ratio operate as an index of both LA and LV pressures. It is therefore likely that the LA and LV remodeling in AF are closely interrelated. However, in a recent cohort of permanent AF with preserved ejection fraction no such analogous changes were found; on the contrary the exercise capacity and E/e’ ratio did not improve by spironolactone, but PALS increased significantly (24). The unresponsiveness to MRA in the latter study which included older patients with more prevalent comorbidities such as diabetes and hypertension, and indices of worse LV dysfunction compared to our cohort, suggests that reversal of structural remodeling in AF might have a point-of-no-return where more severely diseased AF patients, hence severe cardiac remodeling, remain unaffected by MRA. Another possible explanation to the chamber-specific differences in atrial and ventricular responses to spironolactone may reflect different expressions of angiotensin II type 1 receptors and receptor density (25,26).

From a clinical perspective, no between-group differences in incident AF recurrence, DC cardioversions or QOL were detected. Although present study was statistically underpowered to address clinical outcomes, our results are consistent with the overall lack of anti-arrhythmogenic effect of MRA in the setting of AF with preserved ejection fraction (27). Only one small open label trial and a subgroup analysis reported improved sinus rhythm maintenance, symptom status and QOL by spironolactone (28,29). As expected, spironolactone increased serum creatinine and potassium to a greater extent compared to placebo but did not excide adverse events reported earlier and was overall a safe additive to existing background medication.

## LIMITATIONS

Despite the well-balanced randomization and the placebo-controlled interventional design, the small sample size, single-center conduction including only Caucasian ethnicity limit the generalizability to a broad AF population. Another limitation was that the therapeutic effects of MRA were exclusively evaluated by echocardiographically derived cardiac markers that were not corroborated with measures such as serum levels of procollagen peptide fragments or hemodynamic measures during exercise as previously used in related MRA studies. However, in this predominantly healthy population without apparent LV dysfunction, serum biomarkers would likely not have contributed with additional information. Further, it cannot be excluded that the favorable effects of spironolactone presented here might be caused by other effects such as lowering free aldosterone hormone, which was not measured. More importantly, the lack of effect on volume and geometry might be attributed to a low dose of spironolactone combined with a short follow-up, as structural reverse-remodeling quantified with standard echocardiography with current technique and image resolution may reasonably take time and become apparent after long-term therapy.

## CONCLUSION

To conclude, patients with paroxysmal or persistent AF with preserved LVEF treated with spironolactone in conjunction with contemporary AF treatment for 12 months demonstrated improved left atrial reservoir function and left ventricular diastolic properties independent of changes in blood pressure, but did not affect left atrial volumes or geometry, left ventricular global longitudinal strain, quality of life or recurrent episodes of AF.

## Data Availability

The results are published on EudraCT under Trial Identifier ID: 2013-000797-30. All raw data can be provided when necessary.

## Acknowledgements

The authors are grateful to Takeda Pharma, Denmark for providing this trial with the study medication.

## Author contribution

DR managed patient enrollment, data acquisition and analysis, and drafted the manuscript. DR, PLM, MS, and KE contributed to the conception and design of the work. RW contributed to the interrater variability determination. All authors contributed to the interpretation of data, critically revised the final manuscript, and gave their approval for all aspects of work ensuring integrity and accuracy.

## Abbreviations

ACEI: Angiotensin converting enzyme inhibitor
AF: Atrial fibrillation
ARB: Angiotensin II receptor blocker
CI: Confidence interval
ECG: 12-lead electrocardiogram
GLS: Global longitudinal strain
LA: Left atrium
LV: Left ventricle / left ventricular
LVEF: Left ventricular ejection fraction
MRA: Mineralocorticoid receptor antagonist
PACS: Peak atrial contraction longitudinal strain
PALS: Peak atrial longitudinal strain
PSL: Peak systolic longitudinal strain
PSS: Peak systolic longitudinal strain rate
RAAS: Renin-angiotensin II aldosterone system
SR-s: Peak strain rate

## Supplemental material

### Sample size calculations and considerations

The sample size for addressing the primary outcome was based on previous works investigating reverse remodeling of atrial strain by echocardiography in patients undergoing mitral and aorta valve replacement. Assuming a minimum of 10% improvement with spironolactone and applying the expected variance in echocardiographicaly derived PALS previously seen in patients with paroxysmal AF, we anticipated a sample size of 60 per group to show a significant difference (p<0.05) at a power of 90%. To allow for dropouts, the number of patients in each treatment arm was increased to 65 patients per group.

**Supplemental Table S1:** Full list of the inclusion and exclusion criteria

|  |  |
| --- | --- |
| Inclusion criteria | • Adult male or female patients $\geq 18$ years of age.<br>• Paroxysmal AF on one or more occasions detected on 12-lead ECG or Holter-monitoring with AF-episode lasting $\geq 30$ seconds within the last 12 months prior to inclusion, or persistent AF on one or more occasions that has been cardioverted either pharmacologically or electrically within the last 12 months prior to inclusion.<br>• Symptomatic episodes of AF estimated by an EHRA symptom score II-IV.<br>• Women with childbearing potency must use effective contraception (e.g., implants, hormonal depot injections, combined oral contraceptives, intra-uterine devices or vasectomized partner). Men enrolled in this study must agree to use adequate barrier birth control measures. Reliable contraception should be maintained throughout the study and for 30 days after study drug discontinuation.<br>• Written informed consent signed before any study-specific procedure. |
| Exclusion criteria | • Permanent AF. |
|  | • Previous radiofrequency ablation or surgical therapy for AF.<br>• Heart failure (New York Heart Association [NYHA] class >II or/and left ventricular ejection fraction <45%).<br>• Severe coronary artery disease (acute coronary syndrome (ACS) within the last 6 months prior to the screening visit, previous coronary artery bypass graft [CABG], or stable angina with a Canadian Cardiovascular Society [CCS] angina score ≥II or indication for intervention).<br>• Stroke or transient ischemic cerebral attack within the last 6 months prior to the screening visit.<br>• Pregnant women, breastfeeding women, or women of childbearing potential not on adequate birth control.<br>• Presence of severe and hemodynamically significant valvular heart disease.<br>• Chronic liver disease with a Child-Pugh class B or C.<br>• Any disease expected to limit life expectancy to <1 year.<br>• Participation in another clinical trial, either within the last 30 days prior to the screening visit or ongoing.<br>• Addison's disease.<br>• Ongoing therapy with flecainide, propafenone, amiodarone, dronedarone, or sotalol.<br>• Moderate to severe chronic kidney disease, defined as an estimated glomerular filtration rate [eGFR] ≤45 mL/min/1.73 m<sup>2</sup> by the abbreviated Modification of Diet in Renal Disease (MDRD) formula.<br>• Previous intolerance or contradictions to spironolactone.<br>• Patients who are deemed noncompliant with medical treatment.<br>• Mental disorders suspected to interact with study outcomes or any other patient characteristics that may interfere with adherence to the study protocol, such as dementia and substance abuse.<br>• Any surgical or medical condition that in the opinion of the investigator would jeopardize the evaluation of efficacy or safety of the study medication.<br>• Baseline serum potassium ≥5.0 mmol/L or serum sodium <135 mmol/L (one re-assessment of electrolytes is allowed).<br>• Hypotension defined as systolic blood pressure <90 mmHg and/or diastolic blood pressure <50 mmHg. |
|  | • AF associated with an acute reversible condition, i.e., heart surgery, acute infectious disease, untreated hyperthyroidism, etc.<br>• Geographical and social factors complicating follow-up. |

**Supplemental Table S2:**
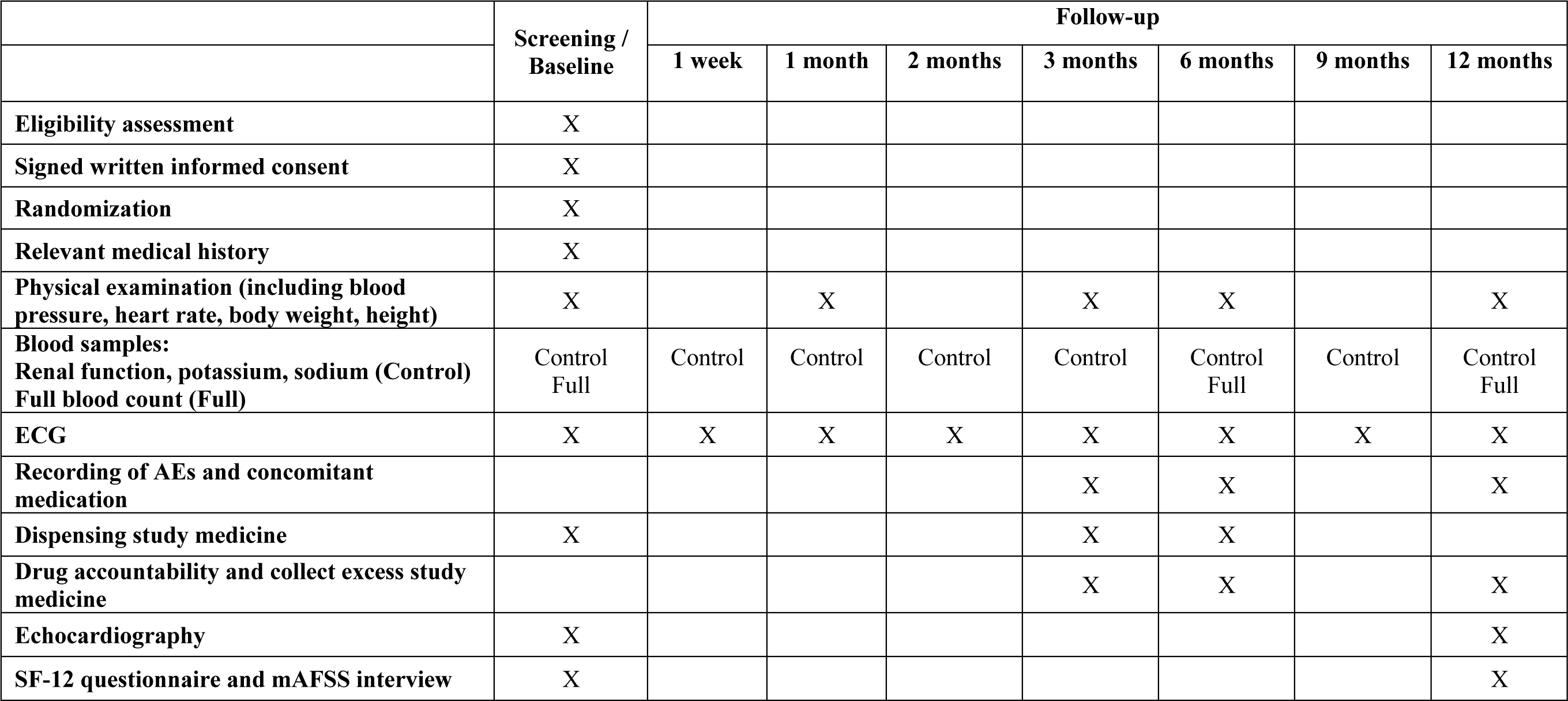
Timeline of study procedures and follow-up assessments

|  | Screening /<br>Baseline | Follow-up |  |  |  |  |  |  |
| --- | --- | --- | --- | --- | --- | --- | --- | --- |
|  |  | 1 week | 1 month | 2 months | 3 months | 6 months | 9 months | 12 months |
| Eligibility assessment | X |  |  |  |  |  |  |  |
| Signed written informed consent | X |  |  |  |  |  |  |  |
| Randomization | X |  |  |  |  |  |  |  |
| Relevant medical history | X |  |  |  |  |  |  |  |
| Physical examination (including blood pressure, heart rate, body weight, height) | X |  | X |  | X | X |  | X |
| Blood samples:<br>Renal function, potassium, sodium (Control)<br>Full blood count (Full) | Control<br>Full | Control | Control | Control | Control | Control<br>Full | Control | Control<br>Full |
| ECG | X | X | X | X | X | X | X | X |
| Recording of AEs and concomitant medication |  |  |  |  | X | X |  | X |
| Dispensing study medicine | X |  |  |  | X | X |  |  |
| Drug accountability and collect excess study medicine |  |  |  |  | X | X |  | X |
| Echocardiography | X |  |  |  |  |  |  | X |
| SF-12 questionnaire and mAFSS interview | X |  |  |  |  |  |  | X |

**Supplemental Table S3:** Withdrawal and/or down-titration of the study medication during follow-up

|  |  |  |
| --- | --- | --- |
| A | No action needed | In case of:<br>Increase in serum creatinine <20% from baseline<br>and/or<br>serum potassium <5.5 mmol/L. |
| B | Temporarily discontinuation of study medication and/or potassium supplements and/or concomitant treatment with ACEI/ARB | In case of:<br>Increase in serum creatinine 20-29% from baseline<br>and/or<br>increase in serum creatinine concentration to $\geq 220$ $\mu\text{mol/L}$<br>and/or<br>mild to moderate hyperkalemia 5.5-6.4 mmol/L |
| C | Withdrawal of the study medication | In case of:<br>Increase in serum creatinine $\geq 30\%$ from baseline or an increase in serum creatinine concentration to $\geq 280$ $\mu\text{mol/L}$<br>and/or<br>severe hyperkalemia ($\geq 6.5$ mmol/L)<br>and/or<br>breast tenderness or discomfort $\geq 1$ month, or visually apparent gynecomastia. |

**Supplemental Table S4:** Intra-observer and inter-observer reliability of echocardiographic parameters

|  | Intra-rater reliability |  | Inter-rater reliability |  |
| --- | --- | --- | --- | --- |
|  | Mean difference<br>(95% CI) | ICC<br>(95% CI) | Mean difference<br>(95% CI) | ICC<br>(95% CI) |
| **LA maximum volume (mL)** | 0.41 (-1.54 to 2.34) | 0.93 (0.92 to 0.95) | -0.83 (-3.58 to 1.91) | 0.86 (0.85 to 0.87) |
| **LA PALS (%)** | -0.21 (-0.90 to 0.49) | 0.88 (0.86 to 0.90) | -0.43 (-1.27 to 0.41) | 0.76 (0.71 to 0.81) |
| **LV GLS (%)** | 0.48 (-0.25 to 1.21) | 0.92 (0.90 to 0.95) | 0.71 (-0.27 to 1.70) | 0.84 (0.80 to 0.90) |

LA indicates left atrium; LoA, limits of agreement; ICC, interclass correlation coefficient; PALS, peak atrial longitudinal strain; LV, left ventricle; EDV, end-diastolic volume; GLS, global longitudinal strain.

**Supplemental Table S5.**
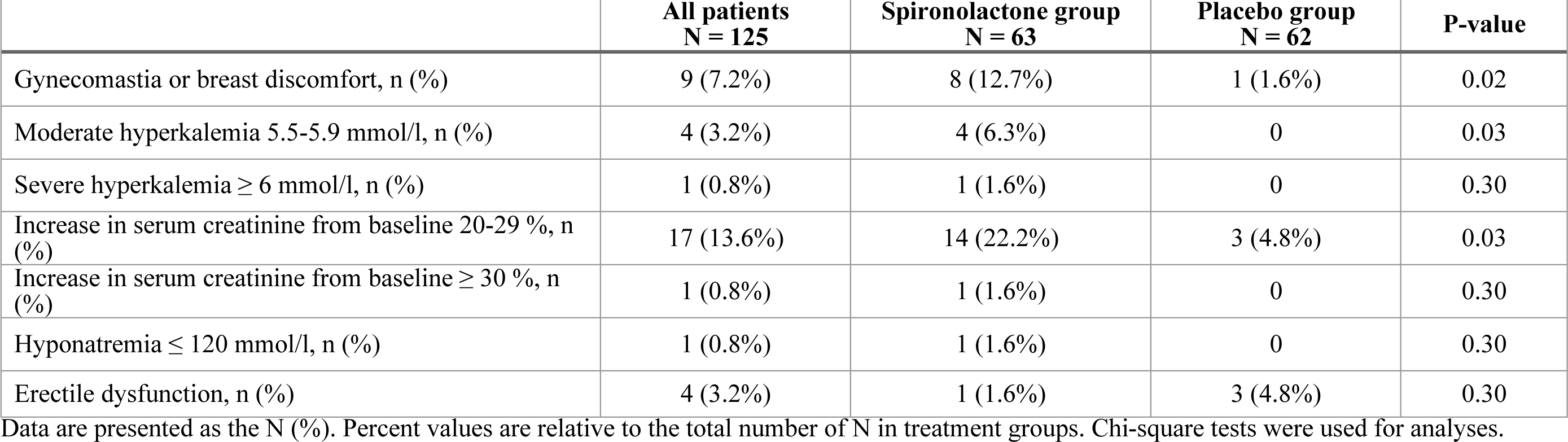
Adverse events of special interest or suspected related to the treatment with spironolactone. Data are presented as the N (%). Percent values are relative to the total number of N in treatment groups. Chi-square tests were used for analyses.

|  | **All patients**<br>**N = 125** | **Spironolactone group**<br>**N = 63** | **Placebo group**<br>**N = 62** | **P-value** |
| --- | --- | --- | --- | --- |
| Gynecomastia or breast discomfort, n (%) | 9 (7.2%) | 8 (12.7%) | 1 (1.6%) | 0.02 |
| Moderate hyperkalemia 5.5-5.9 mmol/l, n (%) | 4 (3.2%) | 4 (6.3%) | 0 | 0.03 |
| Severe hyperkalemia $\geq 6$ mmol/l, n (%) | 1 (0.8%) | 1 (1.6%) | 0 | 0.30 |
| Increase in serum creatinine from baseline 20-29 %, n (%) | 17 (13.6%) | 14 (22.2%) | 3 (4.8%) | 0.03 |
| Increase in serum creatinine from baseline $\geq 30$ %, n (%) | 1 (0.8%) | 1 (1.6%) | 0 | 0.30 |
| Hyponatremia $\leq 120$ mmol/l, n (%) | 1 (0.8%) | 1 (1.6%) | 0 | 0.30 |
| Erectile dysfunction, n (%) | 4 (3.2%) | 1 (1.6%) | 3 (4.8%) | 0.30 |

**Supplemental Table S6.** All registered adverse events and serious adverse events in the study.

|  | **All patients**<br>**N = 125** | **Spirolactone**<br>**N = 63** | **Placebo**<br>**N = 62** | **P-value** |
| --- | --- | --- | --- | --- |
| **Constitutional and neurological:** |  |  |  |  |
| **Fatigue, n (%)** | 30 (24.0%) | 13 (20.6%) | 17 (27.4%) | 0.40 |
| **Headache, n (%)** | 6 (4.8%) | 5 (8.3%) | 1 (1.6%) | 0.10 |
| **Restless legs, n (%)** | 10 (8.0%) | 7 (11.1%) | 3 (4.8%) | 0.20 |
| **Sleep disturbances, n (%)** | 9 (7.2%) | 5 (7.9%) | 4 (6.5%) | 0.70 |
| **Cardiovascular:** |  |  |  |  |
| **Dizziness, bradycardia, orthostatic hypotension, n (%)** | 37 (29.6%) | 25 (39.7%) | 12 (19.4%) | 0.01* |
| **High blood pressure, n (%)** | 14 (11.2%) | 3 (4.7%) | 11 (17.7%) | 0.02* |
| **Chest pain, n (%)** | 13 (10.4%) | 8 (12.7%) | 5 (8%) | 0.40 |
| **Pulmonary:** |  |  |  |  |
| **Dyspnea, n (%)** | 21 (16.8%) | 7 (11.1%) | 14 (22.6%) | 0.01* |
| **Gastrointestinal:** |  |  |  |  |
| **Acid reflux, n (%)** | 14 (11.2%) | 6 (9.5%) | 8 (12.9%) | 0.60 |
| **Abdominal pain, nausea, Bloating, n (%)** | 17 (13.6%) | 9 (14.3%) | 8 (12.9%) | 0.80 |
| **Hematological:** |  |  |  |  |
| **Anemia, n (%)** | 6 (4.8%) | 5 (8.3%) | 1 (1.6%) | 0.10 |
| **Serious adverse events, n (%)** | 4 (3.2%) | 2 (3.2%) | 2 (3.2%) | 0.90 |

**Supplemental Figure S1.**
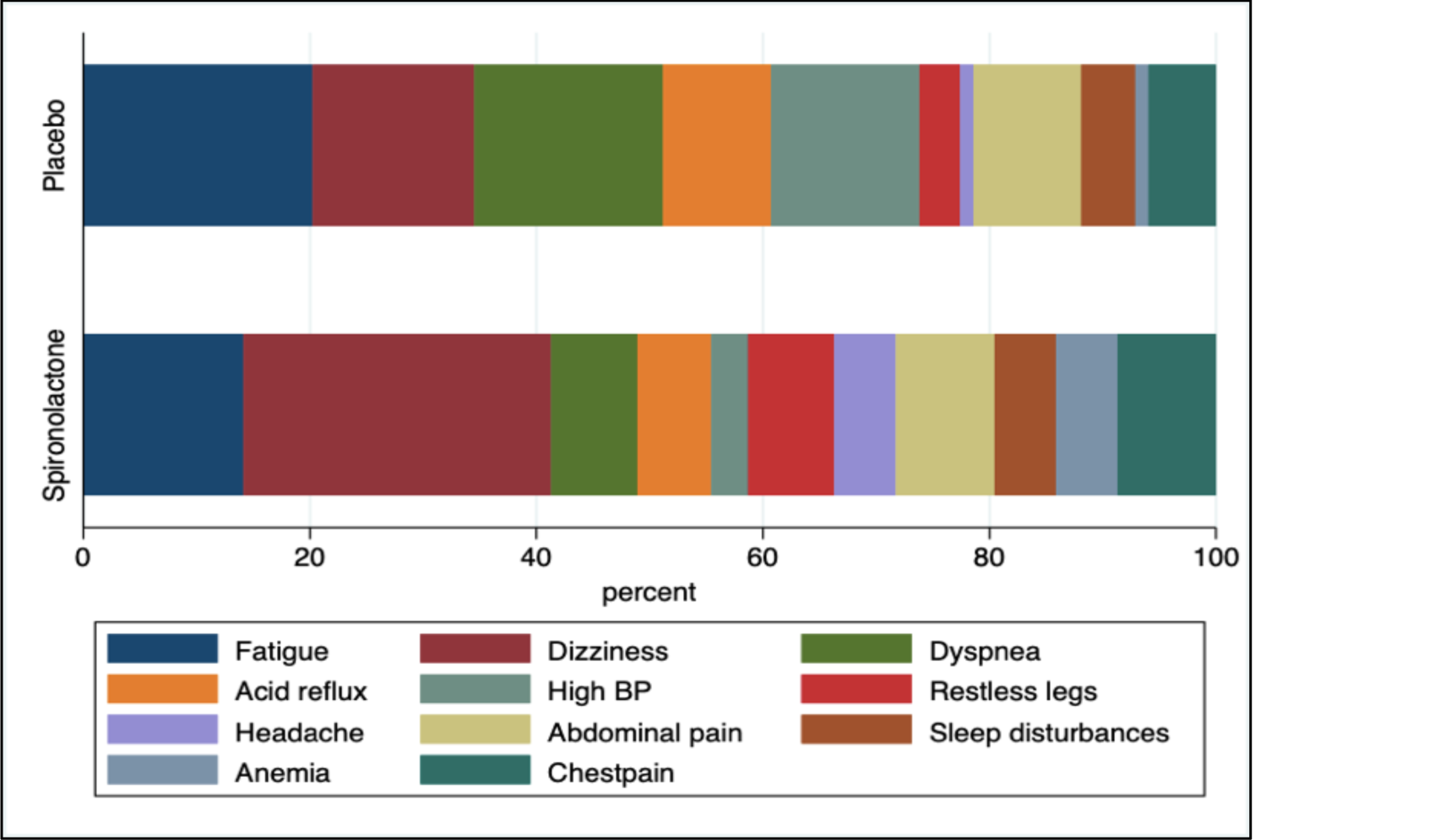
Distribution of proportion of all adverse events by treatment group.

**Table S7.**
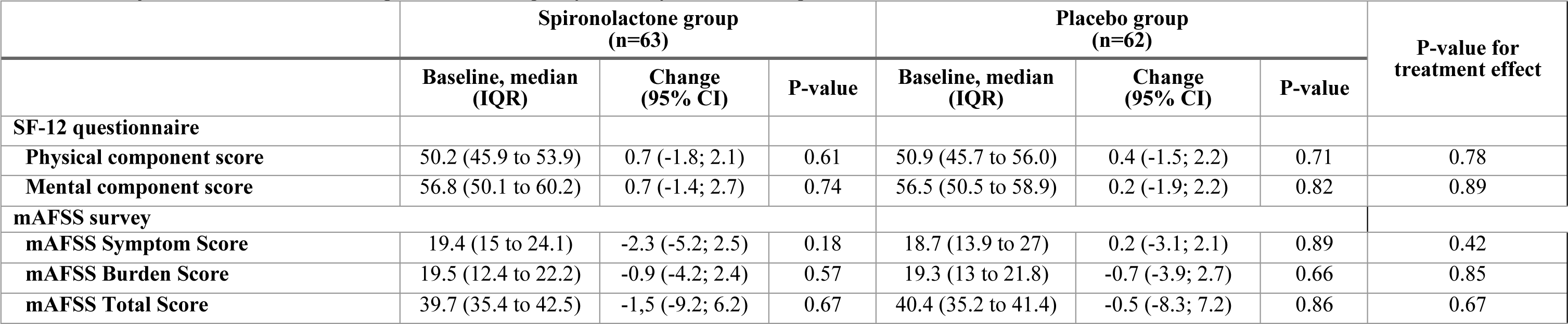
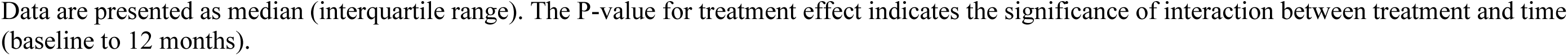
Unadjusted baseline and follow-up health-related quality of life by treatment assignment

|  | Spironolactone group<br>(n=63) |  |  | Placebo group<br>(n=62) |  |  | P-value for<br>treatment effect |
| --- | --- | --- | --- | --- | --- | --- | --- |
|  | Baseline, median<br>(IQR) | Change<br>(95% CI) | P-value | Baseline, median<br>(IQR) | Change<br>(95% CI) | P-value |  |
| SF-12 questionnaire |  |  |  |  |  |  |  |
| Physical component score | 50.2 (45.9 to 53.9) | 0.7 (-1.8; 2.1) | 0.61 | 50.9 (45.7 to 56.0) | 0.4 (-1.5; 2.2) | 0.71 | 0.78 |
| Mental component score | 56.8 (50.1 to 60.2) | 0.7 (-1.4; 2.7) | 0.74 | 56.5 (50.5 to 58.9) | 0.2 (-1.9; 2.2) | 0.82 | 0.89 |
| mAFSS survey |  |  |  |  |  |  |  |
| mAFSS Symptom Score | 19.4 (15 to 24.1) | -2.3 (-5.2; 2.5) | 0.18 | 18.7 (13.9 to 27) | 0.2 (-3.1; 2.1) | 0.89 | 0.42 |
| mAFSS Burden Score | 19.5 (12.4 to 22.2) | -0.9 (-4.2; 2.4) | 0.57 | 19.3 (13 to 21.8) | -0.7 (-3.9; 2.7) | 0.66 | 0.85 |
| mAFSS Total Score | 39.7 (35.4 to 42.5) | -1.5 (-9.2; 6.2) | 0.67 | 40.4 (35.2 to 41.4) | -0.5 (-8.3; 7.2) | 0.86 | 0.67 |

Data are presented as median (interquartile range). The P-value for treatment effect indicates the significance of interaction between treatment and time (baseline to 12 months).

## References

1. Lacolley P, Safar ME, Lucet B, Ledudal K, Labat C, Benetos A. Prevention of aortic and cardiac fibrosis by spironolactone in old normotensive rats. Journal of the American College of Cardiology 2001;37:662–7.

2. Zannad F, Alla F, Dousset B, Perez A, Pitt B. Limitation of excessive extracellular matrix turnover may contribute to survival benefit of spironolactone therapy in patients with congestive heart failure: insights from the randomized aldactone evaluation study (RALES). Rales Investigators. Circulation 2000;102:2700–6.

3. Nattel S, Burstein B, Dobrev D. Atrial remodeling and atrial fibrillation: mechanisms and implications. Circ Arrhythm Electrophysiol 2008;1:62–73.

4. Tsai CT, Chiang FT, Tseng CD et al. Increased expression of mineralocorticoid receptor in human atrial fibrillation and a cellular model of atrial fibrillation. Journal of the American College of Cardiology 2010;55:758–70.

5. Masuda M, Fujita M, Iida O et al. An E/e’ ratio on echocardiography predicts the existence of left atrial low-voltage areas and poor outcomes after catheter ablation for atrial fibrillation. Europace : European pacing, arrhythmias, and cardiac electrophysiology : journal of the working groups on cardiac pacing, arrhythmias, and cardiac cellular electrophysiology of the European Society of Cardiology 2018;20:e60–e68.

6. Cho GY, Marwick TH, Kim HS, Kim MK, Hong KS, Oh DJ. Global 2-dimensional strain as a new prognosticator in patients with heart failure. Journal of the American College of Cardiology 2009;54:618–24.

7. Cameli M, Mandoli GE, Loiacono F, Dini FL, Henein M, Mondillo S. Left atrial strain: a new parameter for assessment of left ventricular filling pressure. Heart Fail Rev 2016;21:65–76.

8. Lang RM, Badano LP, Mor-Avi V et al. Recommendations for cardiac chamber quantification by echocardiography in adults: an update from the American Society of Echocardiography and the European Association of Cardiovascular Imaging. Eur Heart J Cardiovasc Imaging 2015;16:233–70.

9. Aliot E, Botto GL, Crijns HJ, Kirchhof P. Quality of life in patients with atrial fibrillation: how to assess it and how to improve it. Europace : European pacing, arrhythmias, and cardiac electrophysiology : journal of the working groups on cardiac pacing, arrhythmias, and cardiac cellular electrophysiology of the European Society of Cardiology 2014;16:787–96.

10. Dorian P, Jung W, Newman D et al. The impairment of health-related quality of life in patients with intermittent atrial fibrillation: implications for the assessment of investigational therapy. Journal of the American College of Cardiology 2000;36:1303–9.

11. Dorian P, Guerra PG, Kerr CR et al. Validation of a new simple scale to measure symptoms in atrial fibrillation: the Canadian Cardiovascular Society Severity in Atrial Fibrillation scale. Circ Arrhythm Electrophysiol 2009;2:218–24.

12. Shroff SC, Ryu K, Martovitz NL, Hoit BD, Stambler BS. Selective aldosterone blockade suppresses atrial tachyarrhythmias in heart failure. J Cardiovasc Electrophysiol 2006;17:534–41.

13. Kuppahally SS, Akoum N, Burgon NS et al. Left atrial strain and strain rate in patients with paroxysmal and persistent atrial fibrillation: relationship to left atrial structural remodeling detected by delayed-enhancement MRI. Circ Cardiovasc Imaging 2010;3:231–9.

14. Milliez P, Deangelis N, Rucker-Martin C et al. Spironolactone reduces fibrosis of dilated atria during heart failure in rats with myocardial infarction. Eur Heart J 2005;26:2193–9.

15. Zhao J, Li J, Li W et al. Effects of spironolactone on atrial structural remodelling in a canine model of atrial fibrillation produced by prolonged atrial pacing. Br J Pharmacol 2010;159:1584–94.

16. Struthers AD. The clinical implications of aldosterone escape in congestive heart failure. Eur J Heart Fail 2004;6:539–45.

17. Struthers AD. Aldosterone escape during ACE inhibitor therapy in chronic heart failure. Eur Heart J 1995;16 Suppl N:103–6.

18. Pitt B, Zannad F, Remme WJ et al. The effect of spironolactone on morbidity and mortality in patients with severe heart failure. Randomized Aldactone Evaluation Study Investigators. The New England journal of medicine 1999;341:709–17.

19. Izawa H, Murohara T, Nagata K et al. Mineralocorticoid receptor antagonism ameliorates left ventricular diastolic dysfunction and myocardial fibrosis in mildly symptomatic patients with idiopathic dilated cardiomyopathy: a pilot study. Circulation 2005;112:2940–5.

20. Edelmann F, Wachter R, Schmidt AG et al. Effect of spironolactone on diastolic function and exercise capacity in patients with heart failure with preserved ejection fraction: the Aldo-DHF randomized controlled trial. Jama 2013;309:781–91.

21. Kosmala W, Rojek A, Przewlocka-Kosmala M, Wright L, Mysiak A, Marwick TH. Effect of Aldosterone Antagonism on Exercise Tolerance in Heart Failure With Preserved Ejection Fraction. Journal of the American College of Cardiology 2016;68:1823–1834.

22. Kosmala W, Przewlocka-Kosmala M, Szczepanik-Osadnik H, Mysiak A, O’Moore-Sullivan T, Marwick TH. A randomized study of the beneficial effects of aldosterone antagonism on LV function, structure, and fibrosis markers in metabolic syndrome. JACC Cardiovascular imaging 2011;4:1239–49.

23. Mottram PM, Haluska B, Leano R, Cowley D, Stowasser M, Marwick TH. Effect of aldosterone antagonism on myocardial dysfunction in hypertensive patients with diastolic heart failure. Circulation 2004;110:558–65.

24. Shantsila E, Shahid F, Sun Y et al. Spironolactone in Atrial Fibrillation With Preserved Cardiac Fraction: The IMPRESS-AF Trial. J Am Heart Assoc 2020;9:e016239.

25. Burstein B, Libby E, Calderone A, Nattel S. Differential behaviors of atrial versus ventricular fibroblasts: a potential role for platelet-derived growth factor in atrial-ventricular remodeling differences. Circulation 2008;117:1630–41.

26. Pellman J, Lyon RC, Sheikh F. Extracellular matrix remodeling in atrial fibrosis: mechanisms and implications in atrial fibrillation. J Mol Cell Cardiol 2010;48:461–7.

27. Schneider MP, Hua TA, Böhm M, Wachtell K, Kjeldsen SE, Schmieder RE. Prevention of atrial fibrillation by Renin-Angiotensin system inhibition a meta-analysis. Journal of the American College of Cardiology 2010;55:2299–307.

28. Dabrowski R, Borowiec A, Smolis-Bak E et al. Effect of combined spironolactone-β-blocker ± enalapril treatment on occurrence of symptomatic atrial fibrillation episodes in patients with a history of paroxysmal atrial fibrillation (SPIR-AF study). The American journal of cardiology 2010;106:1609–14.

29. De With RR, Rienstra M, Smit MD et al. Targeted therapy of underlying conditions improves quality of life in patients with persistent atrial fibrillation: results of the RACE 3 study. Europace : European pacing, arrhythmias, and cardiac electrophysiology : journal of the working groups on cardiac pacing, arrhythmias, and cardiac cellular electrophysiology of the European Society of Cardiology 2019;21:563–571.

